# Digital thErapy For Improved tiNnitus carE Study (DEFINE): Protocol for a Randomised Controlled Trial

**DOI:** 10.1101/2023.09.25.23296108

**Authors:** Matthew E Smith, Dhiraj Sharma, Oliver Rivero-Arias, Kim Rand, Lucy Barrack, Emma Ogburn, Michael Young, Polly Field, Jan Multmeier, Jameel Muzaffar

## Abstract

Tinnitus is a common health condition, affecting approximately 15% of the UK population. The tinnitus treatment with the strongest evidence base is Cognitive Behavioural Therapy (CBT), with standard tinnitus therapy typically augmented with education, relaxation and other techniques. Availability of CBT and conventional tinnitus therapy more broadly is limited for tinnitus sufferers.

The DEFINE trial aims to assess whether smartphone-delivered tinnitus therapy, the Oto app, is as effective as current standard care, one-to-one therapist-delivered tinnitus treatment for the treatment of tinnitus in adults. The trial is registered in the ISRCTN Registry: ISRCTN99577932.

DEFINE is an open-label, non-inferiority, prospective, parallel design, randomised-controlled trial. Recruitment, interventions and assessments will be remote, enabling UK-wide participant involvement.

198 participants aged 18 years or more will be recruited via social media advertisement or via primary care physicians. A screening process will identify those with tinnitus that impacts health-related quality of life, and following consent smartphone-based audiometry will be performed. Randomisation 1:1 to the Oto app or one-to-one therapist-led tinnitus therapy will be performed centrally by computer, matching groups for age, sex and hearing level. Following participant allocation, the Oto app will be provided for immediate use, or a one-to-one remote therapy appointment booked to occur within approximately 1 week, with up to 6 sessions delivered.

Participant outcomes will be collected at 4,12, 26 and 52 weeks via questionnaire and phone call. The primary outcome is the change in Tinnitus Functional Index (TFI) total score measured at 26 weeks following allocation. Adverse events will be recorded. A health economic evaluation in the form of a cost-utility analysis will be performed using data from participant submitted EuroQol 5D-5L and Health Utilities Index Mark 3 scores and resource use data.

Trial results will be made publicly available, including a plain English summary.

## Introduction

Tinnitus is the perception of sound in the absence of an external stimulus. Tinnitus is very common with approximately 15% of the UK and European population experiencing prolonged tinnitus, and 5-6% considering this bothersome(1). Prevalence varies with age, with approximately 10% of younger adults, 14% of middle-aged adults and 24% of older adults experiencing tinnitus(2). Tinnitus presents a large burden on society: in the UK there are over 1 million GP appointments annually for tinnitus, and the NHS spends c.£750m annually on the condition. Including loss of productivity and other costs, tinnitus is responsible for an annual loss to the economy of £2.7 billion in the UK(3).

Treatment options for people with tinnitus are currently limited. The approach with the strongest evidence base is Cognitive Behavioural Therapy (CBT)(4), with guided self-directed CBT showing the largest effect size and having the highest likelihood of being ranked first in improving tinnitus health-related quality of life (75%), depression (83%), and anxiety (87%)(5). In the UK, current NHS provision as delivered by Ear, Nose and Throat (ENT) surgeons, audiologists and hearing therapists, focuses on ensuring there are no concerning features that may indicate local pathology or systemic health problems. Once these are excluded, National Institute for Health and Care Excellence (NICE) guidance advocates a combination of reassurance, mitigation of any underlying cause if possible, optional hearing aiding if concomitant hearing loss is present, self-care advice and consideration of referral for psychological therapy(6).

Current NHS tinnitus therapy provision is typically based on the principles outlined in Tinnitus Retraining Therapy (TRT), with elements of Cognitive Behavioural Therapy (CBT) and relaxation also incorporated. This provision is typically on a face-to-face, one-to-one basis and lasts between one and ten sessions. Availability of CBT varies, and in the UK a five-fold discrepancy exists in accredited CBT practitioner numbers between the best and least well-served population decile(7). Internet-based CBT has been explored as a therapy for tinnitus(8) and found as effective as face-to-face CBT across different health conditions(9). The proliferation of smartphone technology has led to the development of a number of app-delivered therapies for a variety of conditions, with development accelerated by the COVID-19 pandemic(10). These have been deployed most extensively for mental health conditions with both primary research and evidence synthesis to support their use(11). At present, a number of such apps are available for tinnitus, most utilising sound therapy or CBT(12, 13).

Oto is a multimodal smartphone app-delivered tinnitus treatment programme that combines patient education, CBT, relaxation, mindfulness and a sound library in a customisable package. The self-paced and self-administered delivery method also allows patients to avoid taking time off work and eliminates travel time to a hospital or clinic. In contrast to conventional one-to-one therapy, the smartphone app model removes limits on the number of individuals that can be treated at the same time, and has the potential to significantly improve capacity at a time when waiting lists are under severe pressure.

Many people with tinnitus do not access adequate support, and with a high incidence of the condition and significant cost to individuals and society, there is a need to improve provision to this underserved patient group, as well as supplementing, streamlining and optimising current tinnitus therapy provision.

The DEFINE trial will assess whether smartphone-delivered tinnitus therapy, the Oto app, is as effective as current standard care, one-to-one therapist delivered tinnitus treatment. This will be explored both from the perspective of efficacy to the individual as well as health economic value to the NHS and society more widely.

## Materials and Methods

### Aim, design and setting of the study

DEFINE is an open-label, non-inferiority, prospective, parallel design, randomised-controlled clinical trial. The trial is designed to evaluate the effectiveness of the Oto smartphone tinnitus treatment programme in comparison to one-to-one tinnitus therapy delivered by a trained tinnitus therapist for adults with intrusive tinnitus. Participant outcomes will be assessed over a 12 month period, with the primary outcome being a patient-reported measure of tinnitus severity at 6 months. DEFINE is entirely remotely delivered via a virtual site coordinated by a contract research organisation (Lindus Health), utilising videoconference and electronic technologies for participant recruitment and enrolment, personalised interventions and data capture. With this approach, the trial aims to achieve wide geographical coverage of the United Kingdom. The trial is registered in the ISRCTN Registry: ISRCTN99577932. At time of publication the protocol version is 2.0, dated 7^th^ July 2023.

### Inclusion and exclusion criteria

Adults with tinnitus that has persisted for at least 3 months and is self-assessed as adversely impacting quality of life. Participants must not have previously undergone tinnitus therapy (TRT or CBT by any delivery method).

#### Inclusion Criteria

Participants must meet all the following criteria:

1. Aged 18 years or over
2. Experienced tinnitus symptoms for ≥=3 months, with tinnitus self-assessed as impacting quality of life
3. Have access to a smartphone
4. Able to speak and read English to a sufficient level
5. Able and willing to give consent for the study prior to participation

#### Exclusion criteria

Participants meeting any of the following criteria will not be eligible for inclusion:

1. Tinnitus with potential “red flag” symptoms (e.g., unilateral or pulsatile tinnitus)
2. Significant mental health problems e.g. history of suicidal ideation or requirement for psychiatric/psychological support beyond Primary Care level
3. Have required hospitalisation for mental illness or currently taking an antipsychotic drug
4. Currently taking part in another clinical trial for hearing/tinnitus
5. Awaiting surgical intervention for hearing/tinnitus
6. Previously undergone tinnitus therapy of any type e.g. CBT or TRT

Additional Note: Patients with hearing aids are eligible to participate in the study Pregnant or breastfeeding women (for insurance reasons).

### Sample selection

Potential participants may be identified by:

- Participant self-presentation to the trial website in response to social media (Facebook and other platforms) or patient community advertising. The trial website will host participant information, and participants will be able to register to receive the patient information sheet (Supporting information S4) and a video explaining the trial.
- Clinicians working at invited UK Primary Care practices (Participant Identification Centres - PICs) will describe the study to patients presenting with tinnitus symptoms of at least 3 months duration. If the patient is interested in knowing more about the study they will be directed to the trial website.
- If the above routes do not provide sufficient potential participants, the electronic medical records of patients at participating primary care PICs will be searched by local staff to identify potential participants, who will be invited to take part in the trial via a text message directing them to the trial webpage.

### Randomisation and blinding

Following confirmation of eligibility, the participant will be randomised using a secure, fully-validated and compliant web-based software randomisation system(14).

Participants will be randomised into one of the two intervention arms in a 1:1 ratio, with arms matched via the following minimisation variables and categories:

1. Age (≤40, 41-60, ≥61)
2. Sex (Male, Female)
3. Pure tone average threshold based on screening hearing test (mean threshold at 0.5, 1, 2, 4 kHz (‘None’ <=15dB, ‘Slight’ 16-25dB, ‘Mild’ 26-40dB, ‘Moderate’ 41-55dB, ‘Moderately severe’ 56-70dB, ‘Severe’ 71-90dB, ‘Profound‘ 91dB+)

The trial is open-label, with no blinding of participants or investigators.

### Interventions

#### Oto smartphone tinnitus treatment programme

The Oto app is a digital therapeutic delivered via smartphone. Participants receive personalised tinnitus therapy based on their interaction with the app, with therapy combining CBT with mindfulness, patient education, cognitive exercise and physical therapy (physical stretches/exercises). Participants will be offered a series of daily therapy sessions, where the user listens to recorded audio content that takes them through elements of tinnitus therapy. Participants will work their way through progressive modules using a spiral curriculum(15), where the sessions build on techniques and exercises learned in previous sessions. The programme lasts approximately 6 weeks, using the app 4-5 times per week. Users can personalise their therapy by listening to additional modules and sounds from the sound library. The intervention will begin immediately following randomisation.

#### One-to-one conventional tinnitus therapy

The control arm is designed to represent standard care intervention and was developed based on relevant guidance (BSA Practice Guidance Tinnitus in adults 2021 and NICE guidelines (NG-155)) and via consultation with clinicians providing NHS tinnitus therapy. Participants will undertake one-to-one tinnitus therapy from a trained audiologist / hearing therapist / psychologist via video call.

A detailed specification for the control arm intervention can be found in Supplementary material (S3). In summary, an initial 60 minute session will incorporate a detailed audiological history, an explanation of tinnitus, and therapy tailored to the participant’s needs and understanding, discussing options for treatment and rehabilitation. Questionnaires will be administered as appropriate to assess tinnitus handicap, anxiety and depression in order to establish the functional impact of tinnitus and guide appropriate management. If appropriate, onward referral will be made to other specialties. An individual management plan will then be formulated and agreed with the participant and may include: Relaxation therapy, CBT, Signposting for sound enrichment and relaxation, Recommendation for hearing assessment and/or aid fitting tinnitus maskers where relevant. Signposting will be provided as relevant to trusted resources and local wellbeing services, or to medical care for depression or anxiety.

The control intervention will consist of a minimum of the initial 60 minute tinnitus therapy session, with further 30-60 minute follow up sessions at the discretion of the participant and therapist, up to a maximum of six sessions.

A panel of five therapists will deliver the therapy, selected via interview, and will have training and experience in the treatment of patients with tinnitus using the techniques specified for the control intervention. Participant questionnaires will assess participant perspective of the therapy and will be used for quality assurance.

One-to-one therapy appointments will be offered immediately following randomisation, with the expectation that the first session will occur within 7 days of randomisation. In order to facilitate retention, participants in the control group will be offered access to the Oto App following final data collection at the end of the 12-month trial period.

#### Intervention Adherence

Engagement with the Oto app by participants will be recorded, including frequency of use, overall duration of use and number of modules completed. Engagement with one-to-one therapy will be assessed via therapist reporting, in terms of therapy session dates and number completed for each participant. The number of participants actively using the interventions at 3 and 6 months will be assessed.

At enrolment, participants will be directed to contact their GP for hearing aid referral to a rapid access service such as is provided by accredited ‘Any Qualified Provider’ (AQP) high street audiologists. Any form of hearing aid provision including tinnitus maskers will be permitted and this will be recorded via participant questionnaire at assessments.

No medication is administered within the study, nor are any medications prohibited. Concomitant tinnitus therapy, therapist-led or digital, is not permitted and protocol violations will be screened for at all follow up assessments.

### Assessments

#### Primary outcome

The primary outcome is the difference of the Tinnitus Functional Index (TFI)(16) total score measured at 6 months following allocation and at baseline. The TFI is a 25-item participant-completed questionnaire that is sensitive to changes in tinnitus severity and impact (16), and it is the tool recommended by NICE for this purpose(6). Participants will be administered the questionnaire via the trial’s electronic data capture system using a smartphone or tablet at baseline, and then at 4,12, 26 and 52 weeks.

#### Secondary outcomes

Adverse events will be recorded in both arms from randomisation until and including 12 weeks, captured via participant self-report and structured items in the electronic participant questionnaires.

Two health-related quality of life measures will be used to inform the health economic evaluation: The EuroQol 5D (5 level)(EQ-5D-5L)(17, 18) and Health Utilities Index Mark 3 (HUI3) (19) The EQ-5D-5L is currently the preferred preference-based instrument to measure HRQoL in adults by NICE, and the HUI3 is particularly well-suited to hearing-related research(20). Both will be used as the basis for a health economic assessment (see ‘analyses’).

#### Other assessments

The System Usability Scale (SUS)(21) has become an industry standard for app assessment, and participants in the Oto app intervention arm will be asked to complete this at 12 week follow up. Similarly, the control group will be asked to complete a questionnaire to assess their experience of one-to-one therapy.

### Participant pathway

The participant pathway is summarised in Figure 2.

**Figure 1.**
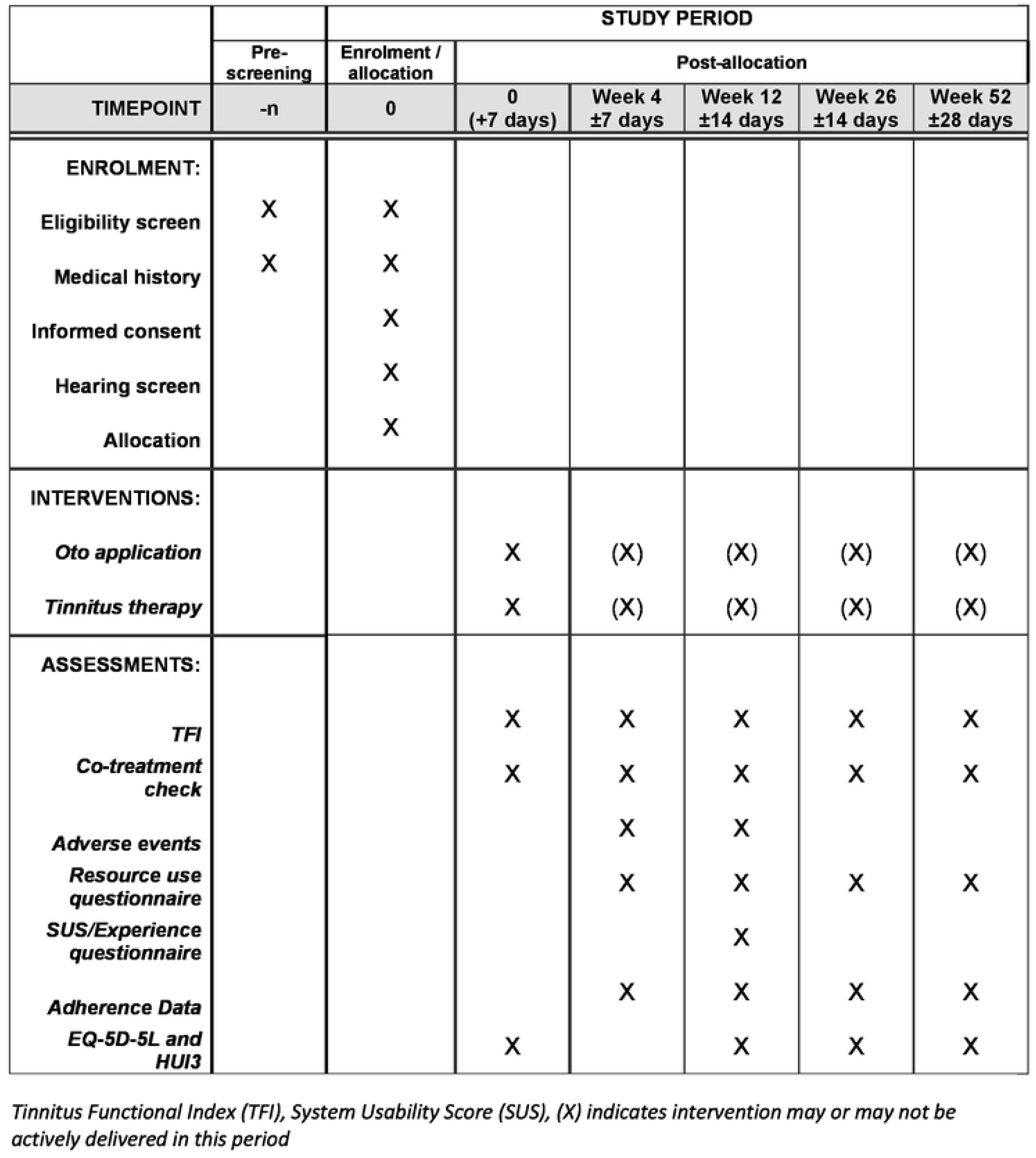
DEFINE trial schedule of enrolment, interventions, and assessments.

**Figure 2.**
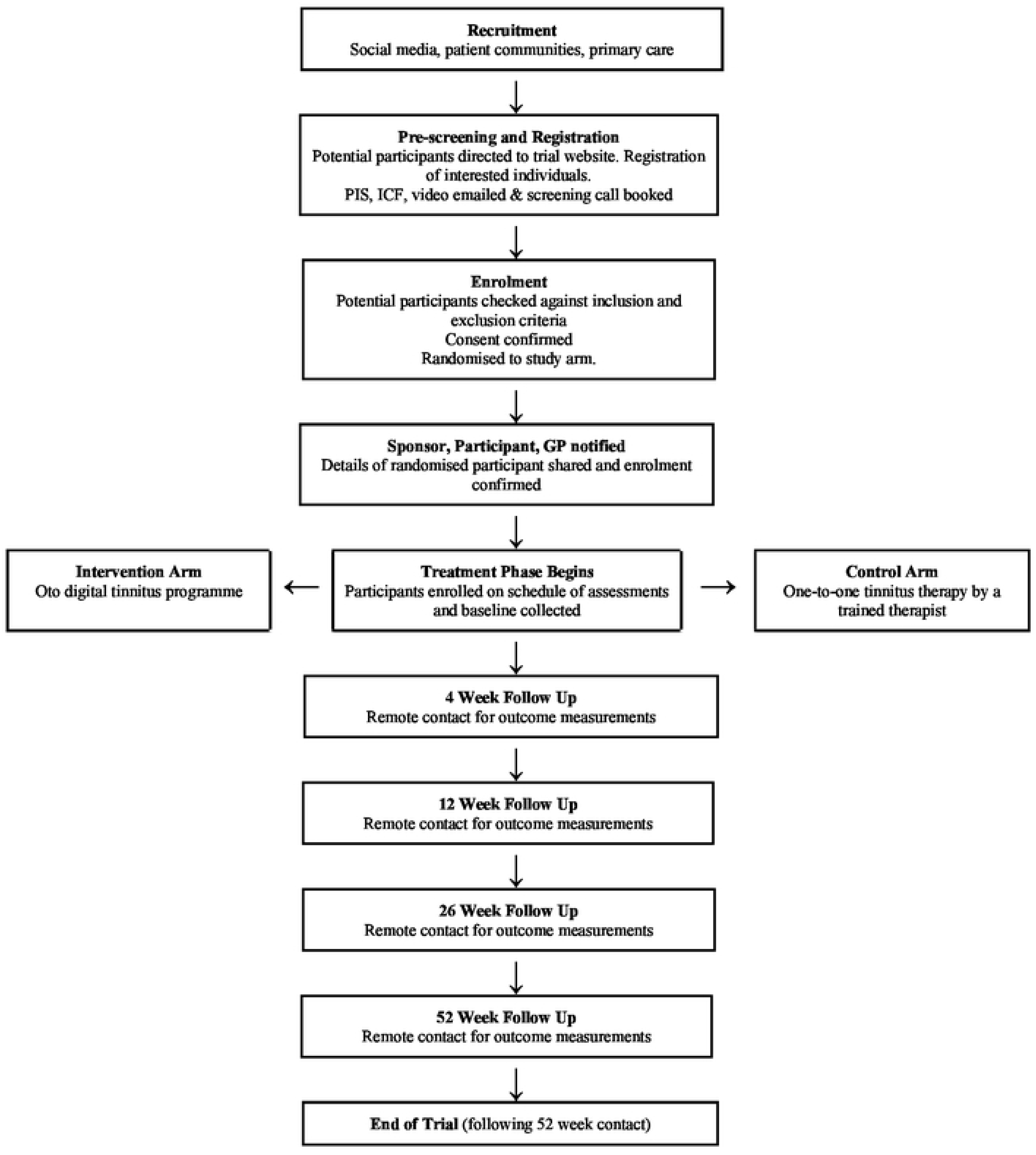

#### Pre-screening

Potential participants will be directed to the trial web page, where they will be informed about the trial aims and design. If interested in taking part, they will complete an online pre-screening form to assess their eligibility for the trial. If potentially eligible, the Patient Information Sheet (PIS) and Informed Consent Form (ICF) will be emailed directly to the participant, as well as a video explaining the trial. If interest is sustained, the potential participant will be asked to book a telephone/video call with a member of the central trial team.

#### Screening and Eligibility Confirmation

Screening will be conducted remotely by a member of the central trial team, during the same telephone/video call where informed consent is obtained. A member of the central trial team will use medical history obtained from the participant to confirm that the individual meets the inclusion and exclusion criteria. A record of screen failures that do not meet the inclusion/exclusion criteria will be retained.

#### Informed consent

Prior to consent, the potential participant will have had the adequate time and opportunity to read all information materials and speak to a suitably qualified and delegated member of the central trial team. If the participant is willing to provide informed consent this will be documented via signature on an electronic consent form (Supporting information S5), and the file will be securely stored. The participant’s general practitioner will be informed of their involvement in the trial.

Identifiable participant details (confidential details, including name and date of birth) will be held in a database separated from the research database after receiving participant consent. The research database will never hold personal identifiable information. Automatic reminders will be sent to participants by doing a one-time recall of the identifiable information by matching the unique study ID.

#### Hearing screening

Participants will be asked to have headphones available at the time of their screening and consent phone call. Participants who consent to trial involvement will be directed to download a smartphone application to screen their hearing for the purpose of stratification at randomisation. Participants with an iOS-based device will be asked to download the ‘Easy Hearing Test’ application (produced by Hiroaki Ito). Those with an Android-based phone will be asked to download the ‘Hearing test’ app (produced by e-audiologica). The participant will then share the hearing test audiogram result with the trial team who will record the mean pure tone threshold (averaged at 0.5, 1, 2 and 4kHz). All participants will be instructed to seek formal audiometric testing via their general practitioner. These screening applications have been validated in the home environment *(publication under peer review)*.

#### Randomisation

A member of the central trial team will inform the participant of their randomisation allocation during their eligibility assessment, and this will be followed up by an email confirmation. Randomisation represents ‘day 0’ of the trial and outcome assessments will be timed from this point.

#### Baseline Measurements-Day 0 (+7 days)

Prior to starting the intervention, participants will report baseline measurements in their survey which will include detailed documentation of tinnitus and audiological history and treatments, as well as completion of trial outcome measures: TFI questionnaire and EQ-5D-5L and HUI3 questionnaires.

#### Administration of interventions

Following baseline data collection, the participant will be granted access to their allocated intervention, with immediate access to the Oto app, or immediate access to booking their first therapist appointment within the following seven days.

#### Follow-up week 4 (±7 days)

The first follow up for data capture will occur at week 4, where participants will be required to complete an online TFI questionnaire. They will be contacted by a member of the trial team to respond to questions about adverse events and any changes in medication or the introduction or modification of hearing devices (e.g. aids).

#### Follow-up week 12 (±14 days)

Further online data capture will occur at week 12, with online TFI questionnaire, SUS and EQ-5D-5L and HUI3 questionnaires. Additional online questionnaires will record any primary and community healthcare appointments and A&E visits related to tinnitus and any personal costs incurred, appointments attended or missed work. A further investigator phone call will cover adverse events and any medication or devices changes.

#### Follow-up week 26 (±14 days) and week 52 (±28 days)

Data capture at 26 weeks will include all data collected at week 12, minus adverse event and SUS data.

#### Data capture

For the duration of the trial, participants will be asked to record their responses to questionnaires directly into the Lindus Health Electronic Data Capture (EDC) platform, via a participant survey link, which will be emailed and/or texted to them.

### Sample size

The present study aims to show that smartphone app-delivered tinnitus therapy is not inferior to CBT in reducing tinnitus symptoms, assessed by the TFI score, after 6 months.

The required sample size was calculated based on a non-inferiority trial design with a non-inferiority margin of 13 points on the TFI scale, as used by two previous studies(22, 23), and a standard deviation of 31 points(24). 13 points corresponds to the minimum clinically important difference for the TFI defined by Meikle *et al.* (16). Assuming a power (1-β) of 80% and a significance level (α) of 2.5%, the sample size was determined as 90 participants per group (180 participants in total). To account for a potential attrition rate of 10%, the total sample size for the study was set at 198 participants.

### Data management plan

Source data will include medical records, hearing screen apps, participant history and questionnaires, app use and therapist-reported data and electronic case report forms.

Data will be entered by both participants and investigators into a validated Electronic Data Capture (EDC) platform hosted on a secure server. Direct access to the EDC platform will be granted to authorised representatives to allow trial-related monitoring and/or audits to ensure compliance with regulations. The data management will be run in accordance with Lindus Health standard procedures, which are fully compliant with Good Clinical Practice (GCP),

GDPR and the Data Protection Act 2018. Research data will be de-identified upon collection. Identifiable information will be stored separately from the research data. A unique trial specific number and/or code in the database will identify the participants.

An online secure data entry system designed to collect sensitive data, such as participant and Study Partner contact details, will be used. All identifiable participant data is encrypted using the Advanced Encryption Standard. Identifiable participant data will be kept for 12 months beyond the end of the study. At the end of the study outcome data will be archived by the University of Cambridge. Non-identifiable participant data will be maintained for a minimum of ten years unless otherwise required to comply with legislation or regulation and reviewed on an annual basis.

### Safety considerations

A Trial Management Group (TMG) is responsible for the day-to-day running of the trial, including monitoring all aspects of the trial and ensuring that the protocol is being adhered to. It will include the Lindus Health Clinical Operations Team, Medical Monitor and Project Manager working closely with the CI and Sponsor and will meet weekly in the first instance. After the initial weekly meetings, the TMG will meet on a regular basis, to discuss study progress and address any issues that may arise.

A detailed plan has been put in place for the detection and management of adverse events. This can be found in the full protocol, included in Supplementary Material S2. Participants will be monitored for adverse events (AEs) through self-report at any point (telephone calls or email), and self-reported via questionnaires, and via direct questioning by trial team members at 4 week follow up call. Trial intervention causality will be assessed by a medical doctor at Lindus Health and/or the CI. AEs judged as being caused by the trial intervention will be reported to the REC that gave favourable opinion, within 15 days of the CI and/or central trial team becoming aware of the event for any AEs judged to be ‘Serious’. AEs will be discussed at the regular TMG meetings and/or referred to the independent Trial Committee for review.

### Analyses

#### Statistical methods

The primary analysis will be conducted on the observed values of the per-protocol population. In addition to this, as a sensitivity analysis a complete-cases, a per-protocol, and an intention-to-treat (ITT) analysis with imputed values will be completed. Compliance will be defined as a minimum of 60 minutes of total use for the Oto app arm, and one or more sessions for the one-to-one therapy group. Participants with poor compliance will be recorded and these participants will be included in the ITT analysis but not the per-protocol analysis.

The primary outcome will be reduction in total TFI score at the 6 month point in the per protocol population. Non-inferiority will be assessed by constructing a 95% confidence interval around the mean difference in TFI reduction between the intervention arms, comparing the upper bound of the confidence interval to the pre-specified non-inferiority margin.

A secondary analysis of the primary outcome will examine significant differences between the control and intervention group’s TFI score at 6 months using a general linear model or a comparable suitable method, using the stratification variables and other potential confounding variables as covariates.

Secondary outcomes will be compared between groups and over time with mixed-effects regression models for repeated measures, using the stratification variables and other potential confounding variables as covariates. Logistic models will be used for binary outcomes, and Gaussian models for continuous outcomes. An interim analysis may be conducted once a sufficient number of participants have completed the 6 month milestone. The results of the interim analysis will not influence the continuation of the trial, and will not be disclosed beyond the trial management group.

#### Handling of Missing and Incomplete Data

The EDC platform provides real-time notifications to the trial team to inform them of any missing data to minimise the risk of missing data. For patient reported outcomes, the EDC will notify the study team if assessments are overdue so the study team can contact the participant up to three times to obtain these data. If a participant reported outcome is not completed within the scheduled window, the data will be considered missing with reasons provided. Missing data will be reported with reasons given where available and imputed using multiple imputation by chained equations (MICE) for the ITT population if more than 10% of TFI change scores cannot be determined due to missing data. The imputed data will be used in a sensitivity analysis to assess the impact of missing data on the primary endpoint.

#### Health economic evaluation

A health economic evaluation forms an integral part of the trial and will be conducted from the perspectives of the NHS, individuals and society. The economic evaluation will take the form of a cost-utility analysis with a one-year time horizon using quality-adjusted life years (QALYs) as the main outcome measure. Although the clinical study has a non-inferiority design and our hypothesis is that QALYs will be similar between the Oto app and standard care, there is no prior hypothesis that costs will be similar between groups. Therefore, a standard economic evaluation framework will be used to synthesise differences in costs and QALYs between study arms using the incremental cost-effectiveness ratio (ICER)(25).

We will follow all participants from baseline to 12 months and will collect information about the delivery of the intervention (app and standard care), primary and community health care, visits to emergency departments and prescribed medications. We will focus on the collection of resource utilisation related to tinnitus. Data will be collected using bespoke questionnaires that will be embedded into the Lindus Health EDC platform for participant completion at regular intervals during follow-up (weeks 12, 26 and 52). Unit costs from recognised national sources including the new NHS National Cost Collection, the Personal and Social Services Research Unit (PSSRU), and the NHS Electronic Drug Tariff will be used to value NHS health care resource use. To understand the burden beyond the health service, we will ask participants about any related out-of-pocket expenses (e.g. hearing aids, complementary and alternative paid treatments) and any time away from paid employment due to the condition.

Tinnitus is known to impair aspects of daily health-related quality of life (HRQoL) and the EQ-5D-5L and the HUI3 will be administered at baseline, and 12, 26 and 52 weeks from randomisation. The health state described in each measure will be converted into a single value on a scale where death is anchored at 0 and perfect health at 1 using value sets. For the EQ-5D-5L the interim 5L-> 3L mapping tool will be used to estimate utility values(26). To calculate utility values from HUI3 responses the Canadian tariff will be used as a UK-specific value set is currently not available. The questionnaires will be administered at baseline and at all follow-up points (weeks 12, 26 and 52). QALYs for each participant over the trial period will be calculated and will be derived by estimating the area under the curve of the profile generated connecting estimates of HRQoL at each time point using linear interpolation. If necessary, any baseline imbalances in HRQoL at baseline and resulting QALYs will be adjusted using regression methods. QALYs will be derived from utility values from the EQ-5D-5L and HUI3 but the former will be used in the base case analysis.

To understand the burden for participants and wider society, healthcare costs will be reported separately from any out-of-pocket expenses and any societal productivity losses. When analysing continuous variables (for example costs, HRQoL scores, QALYs) means and standard deviations will be computed for each trial arm and for comparisons between trial arms, mean differences and 95% parametric confidence intervals will be used. Mean differences in costs and QALYs between study arms will be synthesised using ICERs and to facilitate interpretation, net benefit statistics will be calculated(27). Uncertainty around net-benefits will be expressed using confidence intervals (if feasible) and cost-effectiveness acceptability curves(28). Deterministic sensitivity analysis will be used to address uncertainty around study parameters (e.g. unit costs) not subject to sampling variation. Data completeness on resource use and HRQoL will be closely monitored on a regular basis and missing data mechanisms discussed. Multiple imputation will be implemented to handle missing data if a complete case analysis results in substantial loss of information that could bias our results(29). All analyses will be conducted and reported in accordance with existing good practice guidelines for economic evaluation(30, 31).

### Qualitative Sub-study

A sample of participants will be invited to join focus groups to be conducted approximately 12, 26 and 52 weeks after randomisation. These groups are designed to understand the issues participants may have had accessing tinnitus care before the trial, the way they interacted with the trial interventions, any areas of the interventions that could be improved, and finally to discuss how these interventions may be introduced to the NHS. The focus group findings will be used to aid interpretation of the quantitative study data and the health economic evaluation, and at the end of the trial will be disseminated alongside the main trial results.

#### Sub-study sampling

The DEFINE Trial Manager will generate a list of participants who have consented to further contact for the focus group. Targeted sampling will be conducted 4 months after the start of trial enrolment, and then further rounds will be undertaken 7 and 13 months after first enrolment. Participants will not be selected for more than one focus group timepoint. Where possible the selected sample will be weighted to Office of National Statistics criteria for a nationally representative group based on several variables (age, sex, ethnicity, geographical region), balanced across the randomised intervention given, and the severity of tinnitus (baseline TFI). Consent for focus group participation and data analysis is via a separate consent form for the sub-study.

#### Interview and focus group design

Focus groups and individual participant interviews will be semi-structured, guided by a topic outline. The first set of interviews will be conducted at approximately 12 weeks following allocation, consisting of 1:1 interviews with approximately 10 participants. The second round of participant engagement will occur at 26 weeks following allocation, featuring focus group discussions with 4-5 participants in each group. Focus groups and interviews will be conducted online using video conferencing software. The selection of participants will ensure diversity in age, sex, ethnicity, geographical location, and tinnitus severity. A trained facilitator who is part of the DEFINE trial team will coordinate and guide both the individual and group interviews.

#### Focus group topics

A semi-structured topic guide will be used by the facilitator to initially engage participants in key topics and prompt focussed discussions (Supporting information S6). The guide will explore: barriers to seeking and obtaining advice and treatment for tinnitus; participant ideas, concerns and expectations about the trial interventions; participant interaction with the trial interventions; potential improvements to the delivery or content of the trial interventions; deployment of the interventions within the NHS.

#### Analysis and outcomes

Transcript files will be downloaded from Microsoft Teams and checked against the original video recording for accuracy. Transcripts will be analysed using an inductive thematic analysis, following a 6-step process(32) adapted for group analysis(33). The initial familiarisation phase will involve two investigators jointly reviewing the focus group data to discuss and establish an overall sense of the data. The investigators will then read the transcripts individually before working jointly again to generate codes and themes. The themes will then be reviewed and defined, before producing the report.

### Ethical considerations

The trial will be conducted in accordance with the principles of the Declaration of Helsinki (2013) and with Good Clinical Practice. The trial staff will ensure that the participants’ anonymity is maintained. The study has been granted favourable ethical opinion by the UK Health Research Authority (West-Midlands committee), Ethics Reference: 23/WM/0146, IRAS ID: 328487.

An independent committee has been formed for trial oversight and will meet regularly during the study period. The committee is composed of individuals with expertise in the management of tinnitus, clinical research and statistical analysis. The independent committee will advise the TMG on study design and conduct, and in particular on any interventions that may be required to ensure participant safety or trial viability.

### Status and timeline of the study

The trial commenced recruitment and enrolment on 26/07/2023. It is anticipated that enrolment will be complete by the end of October 2023. Individual participants will have an approximate 12 month period of study involvement.

## Discussion

The DEFINE trial is innovative in the fully-remote nature of its conduct: at no point during recruitment, enrolment, intervention or assessment do investigators and participants need to be co-located. This design underlies many of the trial’s strengths, and also some of its weaknesses. Remote management has enabled very rapid set-up and equitable access to potential participants across the country, removing costs and barriers such as the requirement for travel or in-working hours treatment or assessment. This will aid applicability of the findings to a broad population, hopefully ensuring a diverse cohort. It does however limit participation and treatment to those with a smartphone and sufficient technical ability. At the time of trial reporting, the participant demographic details will be assessed for applicability to the wider UK population. The remote approach also prevents the collection of certain data, such as formal pure-tone audiometry at baseline. While we would not expect audiometry to change with the interventions, it would ideally be available for background data and minimisation purposes. The remote solution used instead of formal audiometry, smartphone-based apps, have been found by our group in another study to have good reliability, and therefore represent an adequate means to match participants in intervention arms.

Ultimately it is hoped that interventions such as the Oto app will enable delivery of tinnitus treatment to a much wider population than those currently treated, possibly including those with less severe symptoms who may not currently be seen in secondary care. For this reason, DEFINE will recruit participants who may not engage with conventional healthcare routes for tinnitus treatment. It is possible that the cohort may not fully represent those currently treated with face-to-face therapy, but this is true of all clinical trials with well-established issues of key groups often being under-served by research(34). Indeed, it is hoped that this and similar remotely-delivered, low participant-burden studies may help to enhance and diversify research inclusion.

The trial is designed to test the effectiveness of the Oto app against standard tinnitus therapy. Standard therapy is difficult to specify, and is highly varied across the UK and internationally, both in terms of availability and content. We have based the control arm intervention on NICE guidance(6) and current UK practice. The control intervention is however not a complete reflection of NHS practice: patients will have rapid access to one-to-one therapy, to ensure outcomes are assessed at comparable timepoints between intervention arms, but the remote delivery setup will also prevent formal audiometric testing by the therapist. Hearing aid provision for rehabilitation of hearing loss is recommended for tinnitus therapy, but in the DEFINE trial this will only be delivered indirectly, via primary care referral of individuals to independent “Any Qualified Provider” (AQP) or NHS audiometry services. Trial participant provision and use of hearing aids will be recorded and analysed. While this aspect of the trial pathway deviates from current holistic best practice, the trial does reflect current service provision in some areas, as well as a possible future management strategy to manage a greater number of affected individuals.

### Dissemination plans

The study results will be published in an open-access journal or repository, ensuring that the research findings are accessible to a wide audience, including healthcare professionals, researchers, and the public. The Chief Investigator will lead the collaborative drafting of the manuscripts, abstracts, press releases and any other publications arising from the study. Authorship will be determined in accordance with the International Committee Medical Journal Editors guidelines and other contributors will be acknowledged. In addition, a plain English summary of results developed in conjunction with the trial Public and Patient Involvement group will be widely distributed, including via patient groups, the trial website, Lindus Health and Oto Health.

## Data Availability

No datasets were generated or analysed during the current study. All relevant data from this study will be made available upon study completion.

## Authors’ contributions

**Matthew E Smith:** Conceptualization, Formal Analysis, Funding Acquisition, Investigation, Methodology, Supervision, Visualisation, Validation, Writing – Original Draft Preparation, Writing – Review & Editing

Dhiraj Sharma: Conceptualization, Data Curation, Formal Analysis, Funding Acquisition, Investigation, Project Administration, Resources, Writing – Original Draft Preparation, Writing – Review & Editing

Oliver Rivero-Arias: Data Curation, Formal Analysis, Funding Acquisition, Investigation, Methodology, Visualisation, Writing – Review & Editing

Kim Rand: Data Curation, Formal Analysis, Funding Acquisition, Investigation, Methodology, Visualisation, Writing – Review & Editing

Lucy Barrack: Data Curation, Formal Analysis, Investigation, Methodology, Project Administration, Resources, Software, Validation, Visualisation, Writing – Review & Editing

Emma Ogburn: Data Curation, Formal Analysis, Funding Acquisition, Investigation, Methodology, Project Administration, Resources, Supervision, Writing – Review & Editing

Michael Young: Conceptualization, Funding Acquisition, Investigation, Methodology, Project Administration, Resources, Writing – Review & Editing

Polly Field: Data Curation, Formal Analysis, Investigation, Methodology, Visualisation, Writing – Review & Editing

Jan Multmeier: Data Curation, Formal Analysis, Funding Acquisition, Investigation, Methodology, Visualisation, Writing – Review & Editing

Jameel Muzaffar: Conceptualization, Formal Analysis, Funding Acquisition, Investigation, Methodology, Supervision, Visualisation, Validation, Writing – Original Draft Preparation, Writing – Review & Editing

## Acknowledgements

The trial sponsor is Oto Health Ltd, 4th Floor, Silverstream House, 45 Fitzroy Street, London, England, W1T 6EB. The Sponsor was involved in study design and will participate in trial delivery. The development of this research was supported by the NIHR Cambridge Biomedical Research Centre. Mrs Rachel Knappett provided expert clinical guidance on control arm design. An expert independent committee is providing oversight and guidance to the trial team: Ass. Prof. Nischay Mehta (Chair), Prof. Simon Lloyd (clinical advisor), Mr Samir Mehta (statistical advisor).

## Supporting Information

S1: SPIRIT schedule

S2: Full protocol

S3: Control arm specification

S4: Patient information sheet

S5: Informed Consent Form

S6: Topic guide

